# The relative impact of vaccination momentum on COVID-19 rates of death in the USA in 2020/2021. The forgotten role of population wellness

**DOI:** 10.1101/2022.03.01.22271721

**Authors:** Victor Keddis

**Affiliations:** University of Melbourne

## Abstract

It is widely accepted that individual underlying health conditions contribute to morbidity and mortality associated with COVID-19; and by inference population wellness will also contribute to COVID-19 outcomes. In addition, over the last two years the predominant pharmaceutical public health response to COVID-19 has been vaccination momentum (i.e. mass and rapid inoculation campaigns).

This paper aims to compare vaccination momentum throughout 2021 and measures of population wellness to estimate the relative impact of each on deaths attributed to COVID-19 across the 50 States of America, plus Washington DC, during 2020 (i.e. the pre-vaccination period) and 2021 (i.e. the vaccination period).

Our analysis shows that: (a) COVID-19 rates of death in 2020 are more important, and statistically more significant, at predicting rates of death in 2021 than vaccination momentum during 2021; (b) vaccination momentum does not predict the magnitude of change in COVID-19 rates of death between 2020 and 2021; and (c) for several underlying heath and risk factors vaccination momentum is significantly less important than population wellness at predicting COVID-19 rates of death.

Of particular interest are our observations that exercise and fruit consumption are 10.1 times more significant at predicting COVID-19 deaths than vaccination momentum, obesity (BMI 30+) is 9.6 times more significant at predicting COVID-19 deaths than vaccination momentum, heart attacks are 4.37 times more significant at predicting COVID-19 deaths than vaccination momentum and smoking is 3.2 times more significant at predicting COVID-19 deaths than vaccination momentum.

If medical and health regulators are to deliver a quantum decrease in COVID-19 deaths they must move beyond the overwhelming focus on COVID-19 vaccination. They must have the courage to urge governments and private organisations to mandate greater exercise, weight loss, less junk food, and better nutrition. And a concerted effort at reducing chronic adverse health conditions.

## Introduction

As early as May 2021 Cummins et al had identified that underlying disease was a significant contributor to COVID-19 morbidity and mortality during the first wave of COVID-19 in the UK [1].

In particular that obesity, type 2 diabetes and chronic kidney disease (CKD) increased the risk of hospitalisation, obesity increased the risk of being admitted to ICU, and underlying CKD, stroke and dementia increased the risk of death.

They also identified the multiplicative effect of multi-morbidity such that, compared to no co-morbidity, the odds ratio of a COVID-19 death is 4.07 times higher for a person with four plus co-morbidities, 2.61 times higher for a person with three co-morbidities, 2.55 times higher for a person with two co-morbidities, and 1.55 times higher for a person with one co-morbidity.

More recently in January 2022, researchers writing in the CDC Morbidity and Mortality Weekly Report found that “Risk for severe outcomes was higher among persons who were aged ≥65 years, were immunosuppressed, or had at least one of eight other underlying conditions” [2]

These eight underlying conditions were immunosuppression, chronic pulmonary disease, chronic liver disease, chronic kidney disease, chronic neurologic disease, diabetes mellitus, chronic cardiac disease, and obesity.

Largely similar findings have been observed by:

a. The Chinese Centre for Disease Control and Prevention which found that cardiovascular disease, hypertension, diabetes, respiratory disease and cancers were associated with an increased risk of death [3];
b. A UK cross-sectional survey of 16,749 patients who were hospitalized with COVID-19 showed that the risk of death was higher for patients with cardiac, pulmonary and kidney disease, as well as cancer, dementia and obesity [4]
c. A French intensive care cohort found that obesity was associated with treatment escalation [5]; and
d. A New York hospital cohort found obesity was associated with hospital presentation [6]

In this paper we extend these observations to a population health perspective by examining for the 50 States of the USA, plus Washington DC, (i.e. 51 Territories):

a. the impact of underlying health and risk factors on selected rates of death (e.g. COVID-19, influenza, pneumonia, all cause, etc);
b. the relative impact of vaccination momentum (2021) and COVID-19 rates of death (2020), on COVID-19 rates of death (2021);
c. the impact of vaccination momentum (2021) on the *change* in COVID-19 rates of death from 2020 to 2021; and
d. the relative impact of vaccination momentum (2021) and underlying health and risk factors on COVID-19 rates of death (2021).

Our approach leverages the variations, between the 51 ‘Territories’, in underlying health and risk factors and vaccination momentum to estimates the impact on rates of death; and *changes* in the rates of death between 2020 and 2021.

## Part A. Data Sources and Definitions

The data sources, definitions, and methodologies used in this paper are detailed below.

### Population Estimates

By Territory, the civilian population by single year of age was extracted from the USA Census Bureau ‘State Population by Characteristics: 2010-2020’ [7]. This data set is based on population *<u>projections</u>* which are updated at each census.

This data set was used to calculate the percentage, of the total Territory population, of each single year of age from 0 to 85 plus. While these percentages are based on population projections, they have remained largely stable over the last 3 to 5 years.

By Territory, the *<u>actual</u>* total civilian population was extracted from the USA Census Bureau ‘National Population Totals and Components of Change: 2020-2021’ [8].

Finally, the *projected* percentage of total Territory population for a single year of age was multiplied by the *actual* total Territory population to arrive at the civilian population by single year of age for 2020 and 2021.

These populations, by single year of age, were subsequently used to calculate rates of deaths and COVID-19 vaccination momentum.

### Underlying Health and Risk Factors

The Behavioural Risk Factor Surveillance System (BRFSS) is the USA’s premier system of health-related telephone surveys that collects state data about U.S. residents regarding their health-related risk behaviours, chronic health conditions, and use of preventive services. [9]

Established in 1984 BRFSS now collects data in all 50 states as well as the District of Columbia. BRFSS completes more than 400,000 adult interviews each year, making it the largest continuously conducted health survey system in the world.

Table 1 gives the underlying health and risk factors extracted from BRFSS for this paper. All factors were extracted as Age Adjusted percentage of Territory population (i.e. age adjusted population prevalence).

**Table 1.**
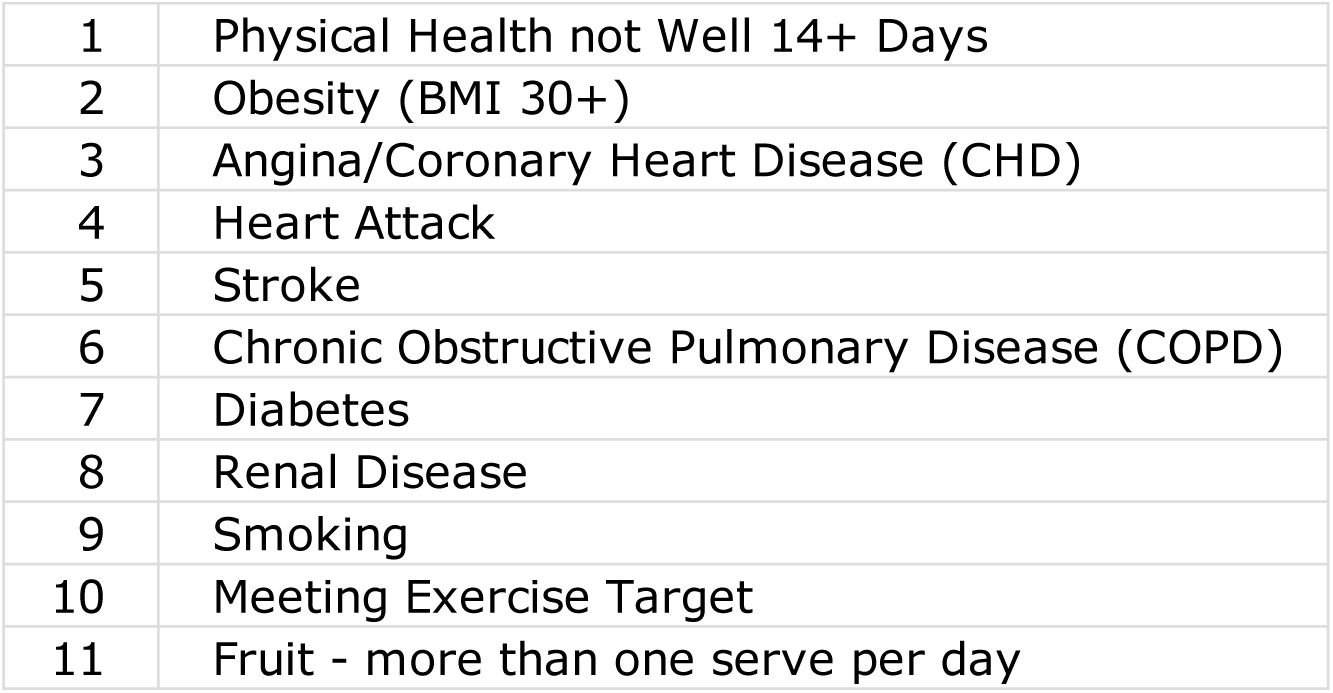
Underlying Health and Risk Factors

### Deaths and Rates of Death – Categories and Definitions

For all 51 Territories, deaths reported to the CDC’s National Centre for Health Statistics (NCHS) was extracted by age group, and Territory of occurrence for 2020 and 2021. [10]

This data set categorised deaths as shown in Table 2, items 1 to 4 and 6 to 7. We extended this data set by calculating two further categories of deaths items 5 and 8.

**Table 2.** Categories of Deaths (Extracted plus Extensions)

| **Table 2.** Categories of Deaths (Extracted plus **Extensions**) |  |  |
| --- | --- | --- |
| **#** | **Code** | **Description** |
| 1 | Covid-19 | Deaths where COVID-19, in addition to other causes, was involved |
| 2 | Pneumonia | Deaths from Pneumonia |
| 3 | Influenza | Deaths from Influenza |
| 4 | Covid-19 and Pneumonia | Deaths where COVID-19 and Pneumonia were jointly involved |
| 5 | Covid Alone | '**Covid Alone**' = Item 1 minus Item 4<br>(i.e. Deaths where COVID-19, in addition to other causes, was involved minus Deaths where COVID-19 and Pneumonia were jointly involved) |
| 6 | Respiratory | Deaths from all respiratory causes (i.e. Pneumonia, Influenza, and COVID-19) |
| 7 | Total | Deaths by all causes |
| 8 | Non-Respiratory | '**Non-Respiratory**' = Item 7 minus Item 6<br>(i.e. Deaths by all causes minus Deaths from all respiratory causes) |

By Territory and age group, ***rates*** of death (per 100k of population) were calculated by simply dividing deaths for each category by the relevant populations in 2020 and 2021; and multiplying by 100,000.

### COVID-19 Vaccination

For the 51 Territories, the daily number of people receiving a dose of vaccine (dose 1, dose 2, and booster) was extracted from the CDC’s vaccination trends tracker. [11]

The data was extracted for all days from 14 December 2020 to 31 January 2022. Together with population data, this daily vaccination data was used to calculate vaccination momentum as described below.

### Vaccination Momentum – Definition and Levels

The purpose of defining and calculating *Vaccination Momentum* is to capture, not only the percentage of a population vaccinated at a given date, but the rate at which that vaccination level is reached.

The theory being that if more people are vaccinated *as quickly as possible* the impact should be lower deaths and lower rates of death.

The first step in calculating Vaccination Momentum was to calculate, by Territory by dose, *Daily Vaccination Weight* for each day between 14 December 2020 and 31 December 2021.

Daily Vaccination Weight is simply the percentage of the population vaccinated on that day multiplied by the number of days from that day to 31 December 2021.

For a given Territory, Vaccination Momentum at 31 December 2021, is then simply the sum of the Daily Vaccination Weights for dose 1 plus the sum of the Daily Vaccination Weights for dose 2 plus the sum of the Daily Vaccination Weights for the booster shot.

By Territory, as at 31 December 2021 Vaccination Momentum is given in Table 3.

**Table 3.** Vaccination Momentum by Terriory by Dose at 31 Dec 2021

| **Table 3** Vaccination Momentum<br>by Territory by Dose at 31 Dec 2021 | Dose 1 | Dose 2 | Booster | Vaccination<br>Momentum | % of Pfizer<br>doses<br>administered | % of Moderna<br>doses<br>administered | % of Janssen<br>doses<br>administered |
| --- | --- | --- | --- | --- | --- | --- | --- |
| Alabama | 150 | 120 | 12 | 281 | 53.3% | 43.8% | 2.9% |
| Alaska | 190 | 161 | 18 | 369 | 57.3% | 38.8% | 4.0% |
| Arizona | 194 | 154 | 15 | 363 | 57.1% | 39.7% | 3.2% |
| Arkansas | 170 | 134 | 14 | 318 | 54.1% | 42.9% | 3.0% |
| California | 211 | 174 | 20 | 405 | 59.1% | 37.7% | 3.2% |
| Colorado | 207 | 176 | 23 | 406 | 57.7% | 38.9% | 3.3% |
| Connecticut | 237 | 199 | 24 | 460 | 58.5% | 38.0% | 3.4% |
| Delaware | 211 | 169 | 19 | 399 | 57.7% | 38.7% | 3.6% |
| District of Columbia | 259 | 187 | 16 | 462 | 61.1% | 36.3% | 2.6% |
| Florida | 203 | 164 | 18 | 384 | 58.7% | 37.2% | 4.1% |
| Georgia | 162 | 128 | 12 | 303 | 58.3% | 39.4% | 2.4% |
| Hawaii | 218 | 189 | 24 | 431 | 60.9% | 36.6% | 2.5% |
| Idaho | 152 | 129 | 15 | 297 | 58.4% | 38.0% | 3.6% |
| Illinois | 194 | 170 | 21 | 385 | 61.0% | 35.9% | 3.2% |
| Indiana | 162 | 141 | 16 | 319 | 60.3% | 36.4% | 3.3% |
| Iowa | 182 | 159 | 22 | 364 | 56.5% | 39.9% | 3.6% |
| Kansas | 184 | 153 | 16 | 353 | 58.7% | 38.2% | 3.1% |
| Kentucky | 169 | 143 | 17 | 329 | 55.0% | 41.3% | 3.7% |
| Louisiana | 159 | 133 | 14 | 305 | 55.6% | 41.1% | 3.2% |
| Maine | 231 | 201 | 27 | 459 | 53.7% | 40.9% | 5.3% |
| Maryland | 212 | 182 | 22 | 416 | 60.3% | 36.5% | 3.2% |
| Massachusetts | 239 | 194 | 23 | 456 | 57.8% | 39.4% | 2.8% |
| Michigan | 175 | 154 | 20 | 349 | 57.8% | 39.1% | 3.1% |
| Minnesota | 200 | 176 | 25 | 401 | 59.8% | 36.5% | 3.6% |
| Mississippi | 151 | 127 | 13 | 291 | 57.1% | 40.5% | 2.4% |
| Missouri | 172 | 141 | 15 | 329 | 60.9% | 36.3% | 2.9% |
| Montana | 175 | 147 | 19 | 341 | 53.8% | 42.3% | 3.9% |
| Nebraska | 184 | 161 | 20 | 365 | 60.7% | 36.3% | 3.0% |
| Nevada | 186 | 146 | 14 | 346 | 61.4% | 34.6% | 4.0% |
| New Hampshire | 232 | 180 | 8 | 420 | 58.2% | 37.4% | 4.4% |
| New Jersey | 208 | 179 | 19 | 406 | 58.5% | 38.0% | 3.5% |
| New Mexico | 213 | 180 | 22 | 415 | 56.0% | 40.8% | 3.2% |
| New York State | 213 | 180 | 18 | 411 | 59.8% | 36.4% | 3.8% |
| North Carolina | 195 | 152 | 10 | 357 | 59.4% | 37.4% | 3.2% |
| North Dakota | 174 | 141 | 16 | 332 | 58.5% | 37.8% | 3.7% |
| Ohio | 172 | 149 | 19 | 340 | 58.2% | 38.2% | 3.6% |
| Oklahoma | 181 | 143 | 14 | 338 | 56.8% | 40.6% | 2.7% |
| Oregon | 205 | 176 | 22 | 404 | 58.8% | 37.4% | 3.7% |
| Pennsylvania | 207 | 168 | 18 | 393 | 57.7% | 38.8% | 3.5% |
| Rhode Island | 224 | 194 | 25 | 443 | 58.7% | 38.2% | 3.1% |
| South Carolina | 170 | 140 | 15 | 325 | 57.6% | 39.2% | 3.2% |
| South Dakota | 191 | 157 | 17 | 365 | 57.7% | 39.3% | 3.0% |
| Tennessee | 164 | 133 | 16 | 313 | 58.7% | 38.5% | 2.8% |
| Texas | 185 | 147 | 14 | 347 | 59.4% | 37.1% | 3.5% |
| Utah | 181 | 148 | 16 | 345 | 60.9% | 35.4% | 3.7% |
| Vermont | 246 | 203 | 32 | 481 | 56.9% | 39.1% | 4.0% |
| Virginia | 212 | 178 | 21 | 411 | 61.0% | 35.8% | 3.2% |
| Washington | 205 | 178 | 22 | 405 | 58.8% | 37.6% | 3.5% |
| West Virginia | 179 | 153 | 19 | 351 | 54.2% | 43.2% | 2.5% |
| Wisconsin | 191 | 166 | 23 | 381 | 58.1% | 38.5% | 3.4% |
| Wyoming | 156 | 132 | 16 | 304 | 52.1% | 44.0% | 3.9% |
| Min | 150 | 120 | 8 | 281 | 52.1% | 34.6% | 2.4% |
| Max | 259 | 203 | 32 | 481 | 61.4% | 44.0% | 5.3% |
| Average | 193 | 161 | 18 | 372 | 58.0% | 38.6% | 3.4% |

## Part B. Understanding COVID-19 Deaths and Survival

In Part B of this paper, for the 51 Territories, we assess:

### Item 1

Impacts of Underlying Health and Risk Factors on Rates of Death;

### Item 2

The relative impact of ‘Vaccination Momentum’ and ‘Covid Alone’ rate of death (2020) on ‘Covid Alone’ rate of death (2021);

### Item 3

Impact of ‘Vaccination Momentum’ on the ***change*** in ‘Covid Alone’ rate of death between 2021 and 2020; and

### Item 4

The relative impact of ‘Vaccination Momentum’ and selected underlying health and risk factors on ‘Covid Alone’ rate of death (2021).

Relative impact will be determined using sum of normalised squared semi-partial correlations derived from multiple-linear regressions. [12]

### Impacts of Underlying Health and Risk Factors on Rates of Death

For the 51 Territories, Table 4 gives the Pearson correlations between selected underlying health and risk factors and rates of death for calendar year 2020.

**Table 4.** Correlations (2020) Rates of Death vs Underlying Health and Risk Factors Figures in red are significant at p < 0.05

| **Table 4.** Correlations (2020)<br>Rates of Death vs Underlying Health and Risk Factors<br>Figures in red are significant at $p < 0.05$ | (1)<br>Death Rate<br>Covid-19 | (2)<br>Death Rate<br>Pneumonia | (3)<br>Death Rate<br>Influenza | (4)<br>Death Rate<br>Covid-19 and<br>Pneumonia | (5)<br>Death Rate<br>Covid Alone | (6)<br>Death Rate<br>Respiratory | (7)<br>Death Rate<br>Total | (8)<br>Death Rate<br>Non-Respiratory |
| --- | --- | --- | --- | --- | --- | --- | --- | --- |
| Physical Health not Well 14+ Days | 0.023009 | 0.229713 | 0.344593 | 0.055388 | -0.013328 | 0.142924 | 0.570732 | 0.634555 |
| BMI 30+ | 0.245443 | 0.287868 | 0.244458 | 0.106088 | 0.297604 | 0.347196 | 0.631488 | 0.627280 |
| Angina/CHD | 0.192621 | 0.381928 | 0.307729 | 0.143395 | 0.178996 | 0.343358 | 0.715361 | 0.730363 |
| Heart Attack | 0.168244 | 0.376617 | 0.295064 | 0.132908 | 0.149394 | 0.325085 | 0.720141 | 0.743383 |
| Stroke | 0.145462 | 0.463088 | 0.238167 | 0.255828 | 0.001320 | 0.297454 | 0.644174 | 0.662329 |
| COPD | 0.116487 | 0.369456 | 0.324106 | 0.100856 | 0.095421 | 0.292321 | 0.775651 | 0.823570 |
| Diabetes | 0.251490 | 0.455446 | 0.398664 | 0.275503 | 0.153608 | 0.374402 | 0.623671 | 0.607046 |
| Renal | 0.156980 | 0.418826 | 0.347059 | 0.258295 | 0.017563 | 0.278765 | 0.544016 | 0.548440 |
| Smoking | 0.145140 | 0.302353 | 0.166010 | 0.068835 | 0.170450 | 0.291677 | 0.754452 | 0.798154 |
| Meeting Exercise Target | -0.327903 | -0.422922 | -0.329962 | -0.199294 | -0.339126 | -0.459244 | -0.596405 | -0.547903 |
| Vegetables - more than one serve per day | -0.416788 | -0.337182 | -0.256831 | -0.393677 | -0.305190 | -0.379562 | -0.069928 | 0.062540 |
| Fruit - more than one serve per day | -0.210991 | -0.388882 | -0.169961 | -0.234480 | -0.122942 | -0.320392 | -0.556414 | -0.553226 |

Correlation significances in columns (2), (3), (6), (7), and (8) are in line with expectations.

However, SARS-CoV-2 being a respiratory virus COVID-19 being largely a respiratory disease [13], correlation significances greater than 0.05 related to COVID-19 rates of death in columns (1), (4), and (5) are unexpected and at odds with the correlation significances in columns (2) and (6).

This may be due to a lack of standardisation, between Territories in 2020, in definitions and recording of COVID-19 deaths.

For the 51 Territories, Table 5 gives the Pearson correlations between selected underlying health and risk factors and rates of death for calendar year 2021.

**Table 5.** Correlations (2021) Rates of Death vs Underlying Health and Risk Factors Figures in red are significant at p < 0.05

| **Table 5.** Correlations (2021)<br>Rates of Death vs Underlying Health and Risk Factors<br>Figures in red are significant at $p < 0.05$ | (1)<br>Death Rate<br>Covid-19 | (2)<br>Death Rate<br>Pneumonia | (3)<br>Death Rate<br>Influenza | (4)<br>Death Rate<br>Covid-19 and<br>Pneumonia | (5)<br>Death Rate<br>Covid Alone | (6)<br>Death Rate<br>Respiratory | (7)<br>Death Rate<br>Total | (8)<br>Death Rate<br>Non-Respiratory |
| --- | --- | --- | --- | --- | --- | --- | --- | --- |
| Physical Health not Well 14+ Days | 0.725489 | 0.679109 | 0.158378 | 0.664119 | 0.597531 | 0.716081 | 0.678733 | 0.594787 |
| BMI 30+ | 0.640051 | 0.536736 | 0.322008 | 0.456577 | 0.713205 | 0.661173 | 0.636523 | 0.561942 |
| Angina/CHD | 0.691397 | 0.668711 | 0.347266 | 0.554104 | 0.682816 | 0.743906 | 0.762689 | 0.692630 |
| Heart Attack | 0.799531 | 0.780166 | 0.311565 | 0.695798 | 0.710443 | 0.832365 | 0.786121 | 0.687699 |
| Stroke | 0.731341 | 0.724827 | 0.517249 | 0.634200 | 0.653095 | 0.771932 | 0.733218 | 0.643185 |
| COPD | 0.724867 | 0.715694 | 0.307401 | 0.591541 | 0.700603 | 0.783055 | 0.842868 | 0.781046 |
| Diabetes | 0.736447 | 0.686984 | 0.503770 | 0.615836 | 0.690441 | 0.759157 | 0.657244 | 0.549688 |
| Renal | 0.719924 | 0.694833 | 0.326190 | 0.642383 | 0.616887 | 0.735430 | 0.612491 | 0.501086 |
| Smoking | 0.675666 | 0.637821 | 0.221333 | 0.525977 | 0.689605 | 0.723772 | 0.808543 | 0.760184 |
| Meeting Exercise Target | -0.503423 | -0.500741 | -0.228099 | -0.348137 | -0.575551 | -0.583621 | -0.564832 | -0.498766 |
| Vegetables - more than one serve per day | -0.284064 | -0.289742 | -0.128752 | -0.329438 | -0.133438 | -0.259165 | -0.005175 | 0.097356 |
| Fruit - more than one serve per day | -0.754796 | -0.711293 | -0.350290 | -0.661828 | -0.661746 | -0.766420 | -0.644045 | -0.528174 |

The unexpected correlation significances (i.e. greater than 0.05) seen in 2020 (refer Table 4 columns (1), (4), and (5)) are now resolved in 2021. This may be attributed to a tighter standardisation, between Territories in 2021 vs 2020, in definitions and recording of COVID-19 deaths.

### Vaccination Momentum and Rates of Death in 2021

To assess the relative impact of Vaccination Momentum in 2021 and previous year’s rates of death (i.e. 2020) on rates of death in 2021, we split the 51 Territories into two groups. These division was based on whether the ‘Covid Alone’ rate of death increased/decreased in 2021 over 2020. There were:

a. 31 Territories (60.8%) where the rate increased, and
b. 20 Territories (39.2%) where the rate decreased.

#### Territories where Rate Increased

For the 31 Territories where the ‘Covid Alone’ rate of death **increased**, Table 6 shows that the largest and most significant predictor (Relative Importance = 98.1%, p=0.0000) of ‘Covid Alone’ death rates in 2021 was ‘Covid Alone’ death rates in 2020.

**Table 6.** Covid Alone. Relative importance of Vaccination Momentum and rate of death (2020) on rate of death (2021) (Adj R^2^=0.7372)

| **Table 6. Covid Alone**<br>Relative importance of Vaccination Momentum and rate of death (2020) on rate of death (2021) (Adj $R^2=0.7372$) | Death Rate 'Covid Alone' 2021 (All Ages)<br>Semi-partial Correlation | Death Rate 'Covid Alone' 2021 (All Ages)<br>p | Relative Importance |
| --- | --- | --- | --- |
| Vaccination Momentum | -0.0810 | 0.3942 | 1.9% |
| Death Rate 'Covid Alone' 2020 | 0.5792 | 0.0000 | 98.1% |

Vaccination Momentum in 2021 played no part in predicting the ‘COVID Alone’ rates of death in 2021.

And for these same Territories, Table 7 shows that the sole and most significant predictor of *all-cause* mortality in 2021 was simply all-cause mortality in 2020.

**Table 7.** All Causes. Relative importance of Vaccination Momentum and rate of death (2020) on rate of death (2021) (Adj R^2^=0.9403)

| **Table 7. All Causes**<br>Relative importance of Vaccination Momentum and rate of death (2020) on rate of death (2021) (Adj R<sup>2</sup>=0.9403) | Death Rate 'All causes' 2021 (All Ages)<br>Semi-partial Correlation | Death Rate 'All Causes' 2021 (All Ages)<br>p | Relative Importance |
| --- | --- | --- | --- |
| Vaccination Momentum | -0.0140 | 0.7566 | 0.0% |
| Death Rate 'All causes' 2020 | 0.9132 | 0.0000 | 100.0% |

Again, Vaccination Momentum played no part in predicting 2021 all-cause mortality.

#### Territories where Rate Decreased

For the 20 Territories where the ‘Covid Alone’ rate of death **decreased**, Table 8 shows that the neither ‘Covid Alone’ rate of deaths in 2020 or Vaccination Momentum were jointly able to predict ‘Covid Alone’ rate of deaths in 2021. Both independent variables were not statistically significant and the whole model adjusted R^2^ was a negligible 0.0661.

**Table 8.** Covid Alone. Relative importance of Vaccination Momentum and rate of death (2020) on rate of death (2021) (Adj R^2^=0.0661)

| **Table 8. Covid Alone**<br>Relative importance of Vaccination Momentum and rate of death (2020) on rate of death (2021) (Adj R<sup>2</sup>=0.0661) | Death Rate 'Covid Alone' 2021 (All Ages)<br>Semi-partial Correlation | Death Rate 'Covid Alone' 2021 (All Ages)<br>p | Relative Importance |
| --- | --- | --- | --- |
| Vaccination Momentum | -0.4050 | 0.0853 |  |
| Death Rate 'Covid Alone' 2020 | 0.1396 | 0.5373 |  |

However, for these same Territories, Table 9 shows that the sole and most significant predictor of all-cause rate of death in 2021 was all-cause rate of death in 2020.

**Table 9.** All Cause. Relative importance of Vaccination Momentum and rate of death (2020) on rate of death (2021) (Adj R^2^=0.8010)

| **Table 9. All Cause**<br>Relative importance of Vaccination Momentum and rate of death (2020) on rate of death (2021) (Adj R<sup>2</sup>=0.8010) | Death Rate 'All causes' 2021 (All Ages)<br>Semi-partial Correlation | Death Rate 'All Causes' 2021 (All Ages)<br>p | Relative Importance |
| --- | --- | --- | --- |
| Vaccination Momentum | -0.1587 | 0.1393 | 5.6% |
| Death Rate 'All causes' 2020 | 0.6496 | 0.0000 | 94.4% |

Vaccination Momentum played a negligible and statistically insignificant part.

### Vaccination Momentum and *Change* in Rates of Death 2021 vs 2020

#### Territories where Rate Increased

For the 31 Territories where the ‘Covid Alone’ rate of death **increased** in 2021 over 2020, Chart 1 shows the *<u>change</u>* in the ‘COVID Alone’ rate of death vs Vaccination Momentum.

**Chart 1.**
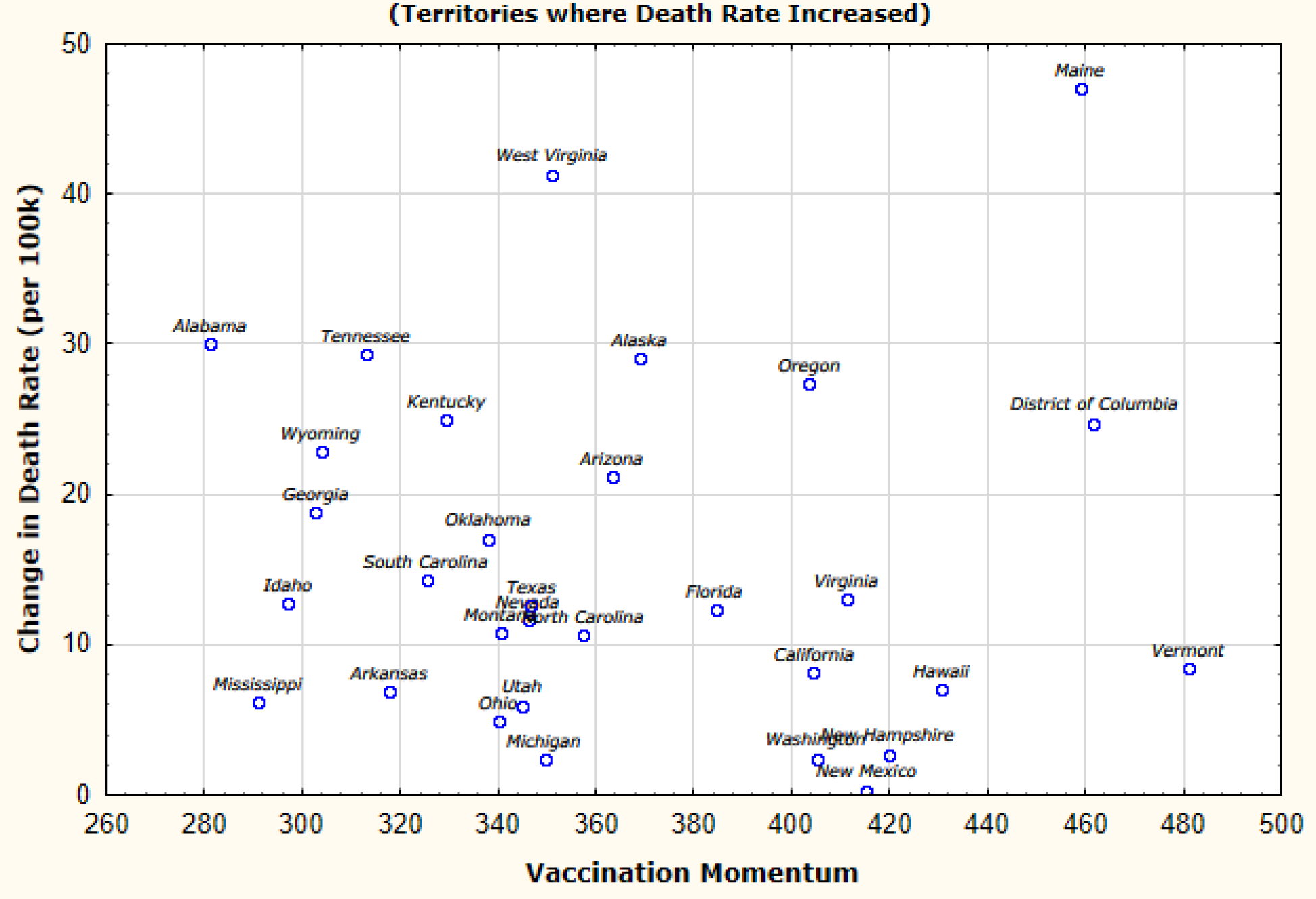
Change in ‘COVID Alone’ Rate of Death 2021/2020 vs Vaccination Momentum.

Table 10 gives the linear regression summary for the data behind Chart 1.

**Table 10.** Covid Alone Rate Increase. Change in death rate vs Vaccination Momentum (Adj R^2^=-0.03367, p=0.8817)

| **Table 10. Covid Alone Rate Increase**<br>Change in death rate vs Vaccination Momentum (Adj R<sup>2</sup>=-0.03367, p=0.8817) | Change in Death Rate (2021/2020) 'Covid Alone' (All Ages)<br>Parameter | Change in Death Rate (2021/2020) 'Covid Alone' (All Ages)<br>p |
| --- | --- | --- |
| Intercept | 17.9421 | 0.2381 |
| Vaccination Momentum | -0.0061 | 0.8817 |

For these 31 Territories, the Spearman Rank Order Correlation between Vaccination Momentum and Change in ‘Covid Alone’ rate of death is −0.1677 (p=0.3670).

Based on Chart 1, Table 10, and the Spearman Rank Order Correlation coefficient, for these 31 Territories, Vaccination Momentum during 2021 played no part in the magnitude of the change in ‘COVID Alone’ rates of death between 2020 and 2021.

#### Territories where Rate Decreased

For the 20 Territories where the ‘Covid Alone’ rate of death **decreased** in 2021 over 2020, Chart 2 shows the *<u>change</u>* in the ‘COVID Alone’ rate of death vs Vaccination Momentum.

**Chart 2.**
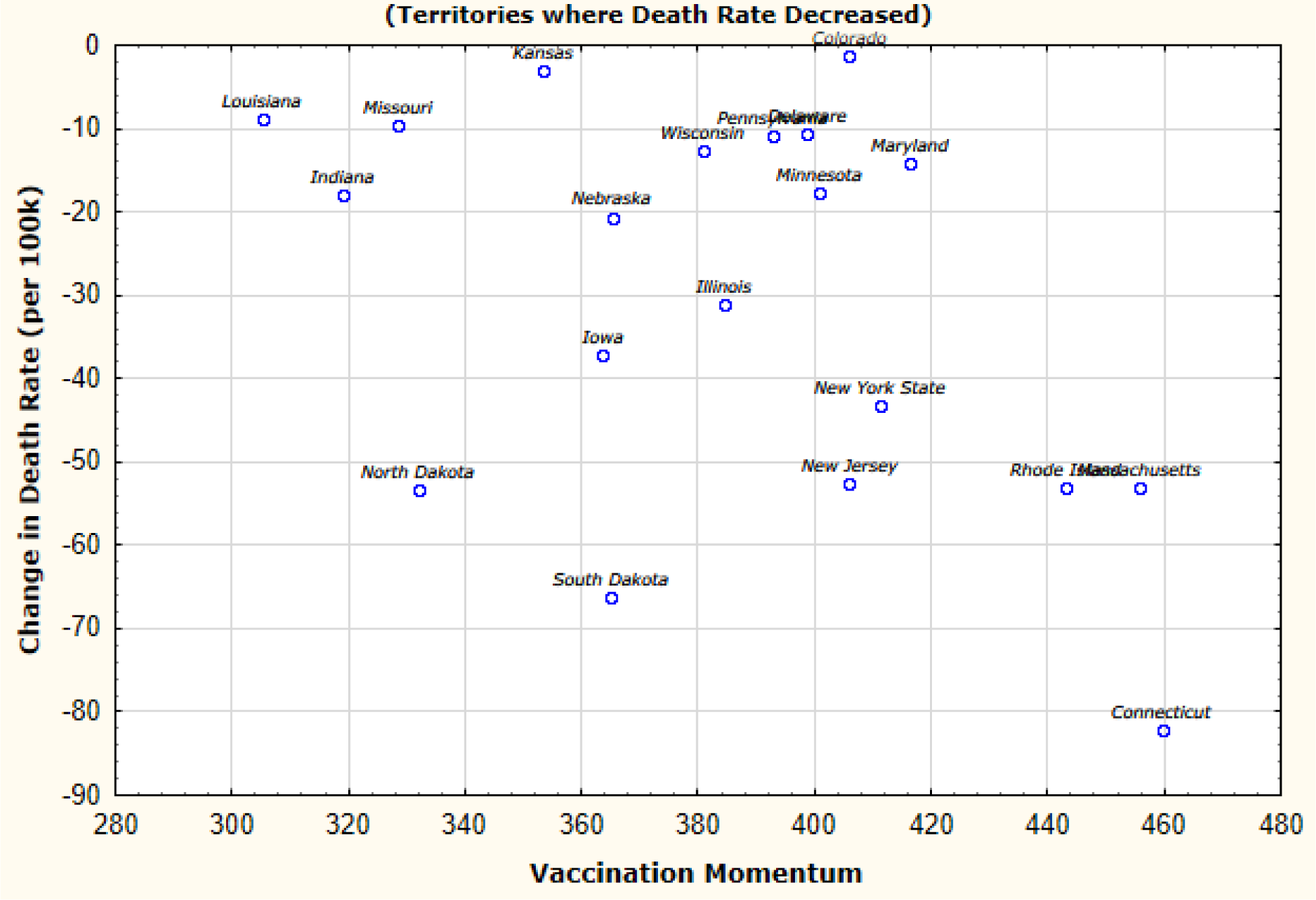
Change in ‘Covid Alone’ rate of death 2021/2020 vs Vaccination Momentum.

Table 11 gives the linear regression summary for the data behind Chart 2.

**Table 11.**
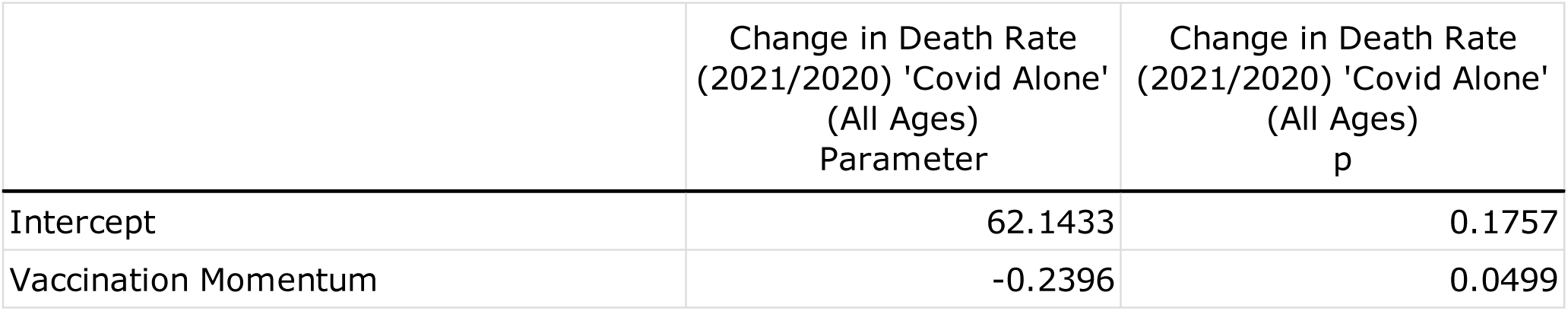
Covid Alone Rate Decrease. Change in death rate vs Vaccination Momentum (Adj R^2^=0.1525, p=0.0498)

| **Table 11. Covid Alone Rate Decrease**<br>Change in death rate vs Vaccination Momentum (Adj R<sup>2</sup>=0.1525, p=0.0498) | Change in Death Rate (2021/2020) 'Covid Alone' (All Ages) Parameter | Change in Death Rate (2021/2020) 'Covid Alone' (All Ages) p |
| --- | --- | --- |
| Intercept | 62.1433 | 0.1757 |
| Vaccination Momentum | -0.2396 | 0.0499 |

For these 20 Territories, the Spearman Rank Order Correlation between Vaccination Momentum and Change in ‘Covid Alone’ rate of death is −0.3714 (p= 0.1068).

Based on Chart 2 and Table 11, for these 20 Territories, Vaccination Momentum during 2021 had a negligible impact on the magnitude of the change in ‘COVID Alone’ rates of death between 2020 and 2021. Vaccination Momentum explained only 15% of the differences in the magnitude of change in ‘COVID Alone’ rates of death between these 20 Territories. And the Spearman Rank Order Correlation was small and statistically insignificant.

### Drivers of ‘Covid Alone’ Rates of Death

In this section we assess the relative impact of Vaccination Momentum vs selected underlying health and risk factors on ‘COVID Alone’ rates of death in 2021.

For the 51 Territories in the data set, relative impact/importance was determined using normalised squared semi-partial correlations derived from multiple-linear regression. [12]

#### Vaccination Momentum vs Physical Health

Table 12 shows the regression summary for ‘Covid Alone’ 2021 as a function of Vaccination Momentum and prevalence of ‘Physical Health not Well 14+ Days’.

**Table 12.** Physical Health. Relative importance of Vaccination Momentum vs Underlying Health or Risk factor on Rate of death ‘Covid Alone’ (2021) (Multiple R^2^=0.4624, p=0.0000)

| **Table 12. Physical Health**<br>Relative importance of Vaccination Momentum vs Underlying Health or Risk factor on Rate of death 'Covid Alone' (2021) (Multiple $R^2=0.4624$, $p=0.0000$) | Death Rate<br>'Covid Alone' 2021<br>(All Ages)<br>Semi-partial<br>Correlation | Death Rate<br>'Covid Alone'<br>2021<br>(All Ages)<br>p | Relative<br>Importance |
| --- | --- | --- | --- |
| Vaccination Momentum | -0.3246 | 0.0035 | 50.4% |
| Physical Health not Well 14+ Days | 0.3218 | 0.0038 | 49.6% |

#### Vaccination Momentum vs Obesity

Table 13 shows the regression summary for ‘Covid Alone’ 2021 as a function of Vaccination Momentum and prevalence of ‘Obesity (BMI 30+)’.

**Table 13.** Obesity. Relative importance of Vaccination Momentum vs Underlying Health or Risk factor on Rate of death ‘Covid Alone’ (2021) (Multiple R^2^=0.5259, p=0.0000)

| **Table 13. Obesity**<br>Relative importance of Vaccination Momentum vs Underlying Health or Risk factor on Rate of death 'Covid Alone' (2021) (Multiple $R^2=0.5259$, $p=0.0000$) | Death Rate<br>'Covid Alone' 2021<br>(All Ages)<br>Semi-partial<br>Correlation | Death Rate<br>'Covid Alone'<br>2021<br>(All Ages)<br>p | Relative<br>Importance |
| --- | --- | --- | --- |
| Vaccination Momentum | -0.1315 | 0.1919 | 9.4% |
| Obesity (BMI 30+) | 0.4088 | 0.0002 | 90.6% |

#### Vaccination Momentum vs Angina/CHD

Table 14 shows the regression summary for ‘Covid Alone’ 2021 as a function of Vaccination Momentum and prevalence of ‘Angina/CHD’.

**Table 14.**
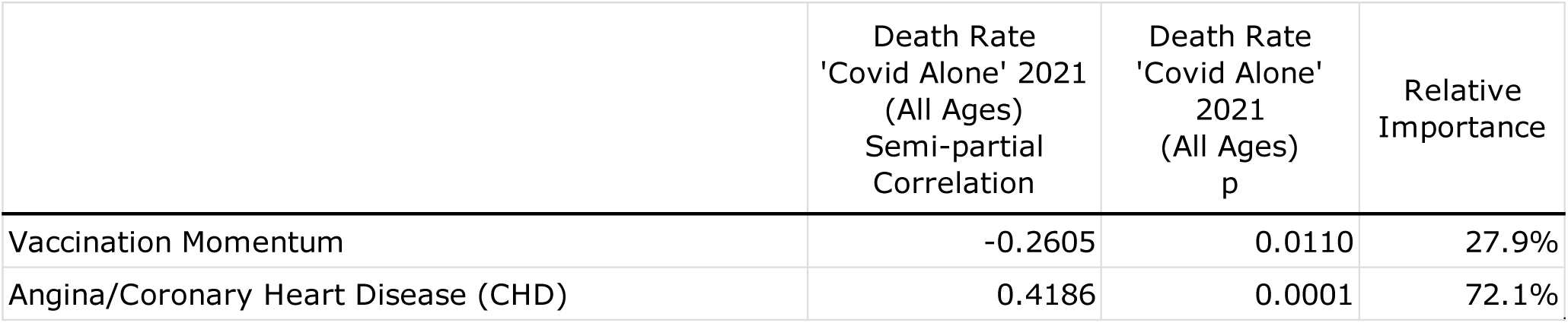
Angina/CHD. Relative importance of Vaccination Momentum vs Underlying Health or Risk factor on Rate of death ‘Covid Alone’ (2021) (Multiple R^2^=0.5341, p=0.0000)

#### Vaccination Momentum vs Heart Attack

Table 15 shows the regression summary for ‘Covid Alone’ 2021 as a function of Vaccination Momentum and prevalence of ‘Heart Attack’.

**Table 15.** Heart Attack. Relative importance of Vaccination Momentum vs Underlying Health or Risk factor on Rate of death ‘Covid Alone’ (2021) (Multiple R^2^=0.5480, p=0.0000)

| **Table 15. Heart Attack**<br>Relative importance of Vaccination Momentum vs Underlying Health or Risk factor on Rate of death 'Covid Alone' (2021) (Multiple $R^2=0.5480$, $p=0.0000$) | Death Rate<br>'Covid Alone' 2021<br>(All Ages)<br>Semi-partial<br>Correlation | Death Rate<br>'Covid Alone'<br>2021<br>(All Ages)<br>p | Relative<br>Importance |
| --- | --- | --- | --- |
| Vaccination Momentum | -0.2081 | 0.0371 | 18.6% |
| Heart Attack | 0.4349 | 0.0000 | 81.4% |

#### Vaccination Momentum vs Stroke

Table 16 shows the regression summary for ‘Covid Alone’ 2021 as a function of Vaccination Momentum and prevalence of ‘Stroke’.

**Table 16.** Stroke. Relative importance of Vaccination Momentum vs Underlying Health or Risk factor on Rate of death ‘Covid Alone’ (2021) (Multiple R^2^=0.4791, p=0.0000)

| **Table 16. Stroke**<br>Relative importance of Vaccination Momentum vs Underlying Health or Risk factor on Rate of death 'Covid Alone' (2021) (Multiple $R^2=0.4791$, $p=0.0000$) | Death Rate<br>'Covid Alone' 2021<br>(All Ages)<br>Semi-partial<br>Correlation | Death Rate<br>'Covid Alone'<br>2021<br>(All Ages)<br>p | Relative<br>Importance |
| --- | --- | --- | --- |
| Vaccination Momentum | -0.2293 | 0.0326 | 30.4% |
| Stroke | 0.3467 | 0.0017 | 69.6% |

#### Vaccination Momentum vs COPD

Table 17 shows the regression summary for ‘Covid Alone’ 2021 as a function of Vaccination Momentum and prevalence of ‘COPD’.

**Table 17.**
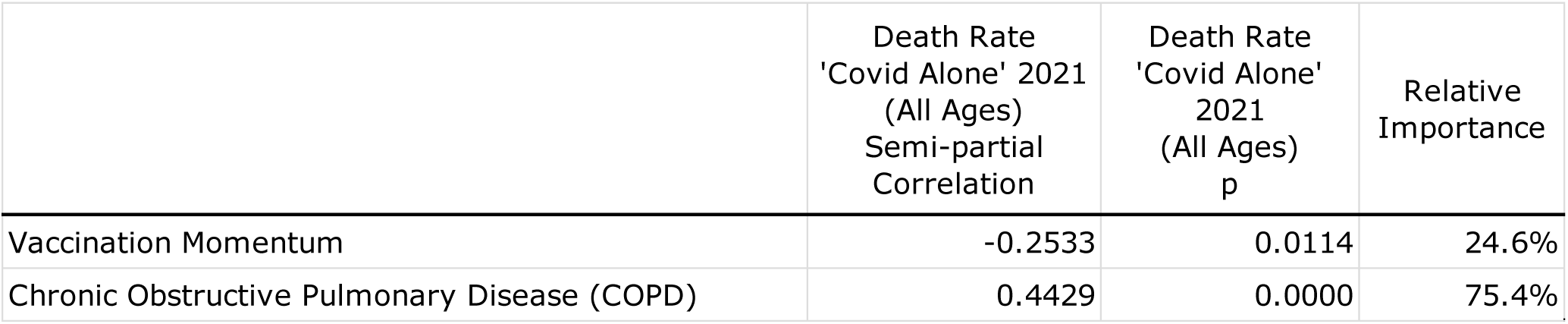
COPD. Relative importance of Vaccination Momentum vs Underlying Health or Risk factor on Rate of death ‘Covid Alone’ (2021) (Multiple R^2^=0.5550, p=0.0000)

| **Table 17. COPD**<br>Relative importance of Vaccination Momentum vs Underlying Health or Risk factor on Rate of death 'Covid Alone' (2021) (Multiple $R^2=0.5550$, $p=0.0000$) | Death Rate<br>'Covid Alone' 2021<br>(All Ages)<br>Semi-partial<br>Correlation | Death Rate<br>'Covid Alone'<br>2021<br>(All Ages)<br>p | Relative<br>Importance |
| --- | --- | --- | --- |
| Vaccination Momentum | -0.2533 | 0.0114 | 24.6% |
| Chronic Obstructive Pulmonary Disease (COPD) | 0.4429 | 0.0000 | 75.4% |

#### Vaccination Momentum vs Diabetes

Table 18 shows the regression summary for ‘Covid Alone’ 2021 as a function of Vaccination Momentum and prevalence of ‘Diabetes’.

**Table 18.** Diabetes. Relative importance of Vaccination Momentum vs Underlying Health or Risk factor on Rate of death ‘Covid Alone’ (2021) (Multiple R^2^=0.5387, p=0.0000)

| **Table 18. Diabetes**<br>Relative importance of Vaccination Momentum vs Underlying Health or Risk factor on Rate of death 'Covid Alone' (2021) (Multiple $R^2=0.5387$, $p=0.0000$) | Death Rate<br>'Covid Alone' 2021<br>(All Ages)<br>Semi-partial<br>Correlation | Death Rate<br>'Covid Alone'<br>2021<br>(All Ages)<br>p | Relative<br>Importance |
| --- | --- | --- | --- |
| Vaccination Momentum | -0.2491 | 0.0144 | 25.6% |
| Diabetes | 0.4241 | 0.0001 | 74.4% |

#### Vaccination Momentum vs Renal Disease

Table 19 shows the regression summary for ‘Covid Alone’ 2021 as a function of Vaccination Momentum and prevalence of ‘Renal Disease’.

**Table 19.** Renal Disease. Relative importance of Vaccination Momentum vs Underlying Health or Risk factor on Rate of death ‘Covid Alone’ (2021) (Multiple R^2^=0.4768, p=0.0000)

| **Table 19. Renal Disease**<br>Relative importance of Vaccination Momentum vs Underlying Health or Risk factor on Rate of death 'Covid Alone' (2021) (Multiple $R^2=0.4768$, $p=0.0000$) | Death Rate<br>'Covid Alone' 2021<br>(All Ages)<br>Semi-partial<br>Correlation | Death Rate<br>'Covid Alone'<br>2021<br>(All Ages)<br>p | Relative<br>Importance |
| --- | --- | --- | --- |
| Vaccination Momentum | -0.3103 | 0.0046 | 44.9% |
| Renal Disease | 0.3434 | 0.0019 | 55.1% |

#### Vaccination Momentum vs Smoking

Table 20 shows the regression summary for ‘Covid Alone’ 2021 as a function of Vaccination Momentum and prevalence of ‘Smoking’.

**Table 20.** Smoking. Relative importance of Vaccination Momentum vs Underlying Health or Risk factor on Rate of death ‘Covid Alone’ (2021) (Multiple R^2^=0.5282, p=0.0000)

| **Table 20. Smoking**<br>Relative importance of Vaccination Momentum vs Underlying Health or Risk factor on Rate of death 'Covid Alone' (2021) (Multiple $R^2=0.5282$, $p=0.0000$) | Death Rate<br>'Covid Alone' 2021<br>(All Ages)<br>Semi-partial<br>Correlation | Death Rate<br>'Covid Alone'<br>2021<br>(All Ages)<br>p | Relative<br>Importance |
| --- | --- | --- | --- |
| Vaccination Momentum | -0.2293 | 0.0250 | 23.7% |
| Smoking | 0.4114 | 0.0001 | 76.3% |

#### Vaccination Momentum vs Exercise and Diet

Table 21 shows the regression summary for ‘Covid Alone’ 2021 as a function of Vaccination Momentum, and prevalence of ‘Exercise’ and ‘Fruit Consumption’ targets.

**Table 21.** Exercise and Fruit Consumption. Relative importance of Vaccination Momentum vs Underlying Health or Risk factor on Rate of death ‘Covid Alone’ (2021) (Multiple R^2^=0.4654, p=0.0000)

| **Table 21. Exercise and Fruit Consumption**<br>Relative importance of Vaccination Momentum vs Underlying Health or Risk factor on Rate of death 'Covid Alone' (2021) (Multiple $R^2=0.4654$, $p=0.0000$) | Death Rate<br>'Covid Alone' 2021<br>(All Ages)<br>Semi-partial<br>Correlation | Death Rate<br>'Covid Alone'<br>2021<br>(All Ages)<br>p | Relative<br>Importance |
| --- | --- | --- | --- |
| Vaccination Momentum | 0.1281 | 0.2408 | 9.0% |
| Meeting Exercise Target | -0.1959 | 0.0757 | 21.0% |
| Fruit - more than one serve per day | -0.3578 | 0.0018 | 70.0% |

## Part C. Discussion

It is now widely accepted that underlying adverse health conditions contribute to morbidity and mortality associated with COVID-19. [1] to [6] And to address COVID-19 morbidity and mortality, the predominate focus has been on mass and rapid inoculation of large percentages of country populations. [14]

However, the relative *impact and contribution* of population wellness to minimising COVID-19 morbidity and mortality has largely been neglected. This paper aims to address this neglect, by studying the relative impact of vaccination momentum versus population wellness across the 50 States of America, plus Washington DC (i.e. 51 Territories), during 2020 and 2021.

Our first observation (refer Table 5) was that underlying health factors (such as prevalence of angina, heart attacks, stroke, COPD, diabetes, and renal disease) are significantly associated with rates of death across all categories of causes. Furthermore, risk factors such as obesity, smoking, insufficient exercise, and poor nutrition were also significantly associated with rates of death across all categories of causes.

Our second observation was that there were 31 Territories where the rate of ‘Covid Alone’ deaths (i.e. deaths attributed to COVID-19 less those deaths where pneumonia was a factor) increased in 2021 over 2020; and 20 Territories where the rate decreased.

For the 31 Territories where the rate increased, vaccination momentum during 2021 played no part in predicting ‘Covid Alone’ rates of deaths in 2021. The largest and most significant predictor (i.e. relative importance = 98%, p=0.0000) was simply ‘Covid Alone’ rates of death in 2020. Furthermore, vaccination momentum played no part in predicting all-cause mortality in 2021.

For the 20 Territories where the rate decreased, neither ‘Covid Alone’ rate of deaths in 2020 or Vaccination Momentum during 2021 were jointly able to predict ‘Covid Alone’ rate of deaths in 2021. However, for these same 20 Territories the sole and most significant predictor of all-cause rate of death in 2021 was all-cause rate of death in 2020. And once Vaccination Momentum during 2021 played a negligible and statistically insignificant part.

Our third observation was that, for the 31 Territories where the ‘Covid Alone’ rate increased, vaccination momentum during 2021 played no part in the magnitude of change between 2021 and 2020. And, for the 20 Territories where the ‘Covid Alone’ rate decreased, Vaccination Momentum explained only 15% of the differences in the magnitude of change in ‘COVID Alone’ rates of death between these 20 Territories.

Our fourth observation was that across all factors of underlying risk and health vaccination momentum during 2021 played a lesser role in predicting ‘Covid Alone’ rates of death in 2021. Table 22 shows the relative importance of selected underlying health and risk factors over vaccination momentum.

**Table 22.**
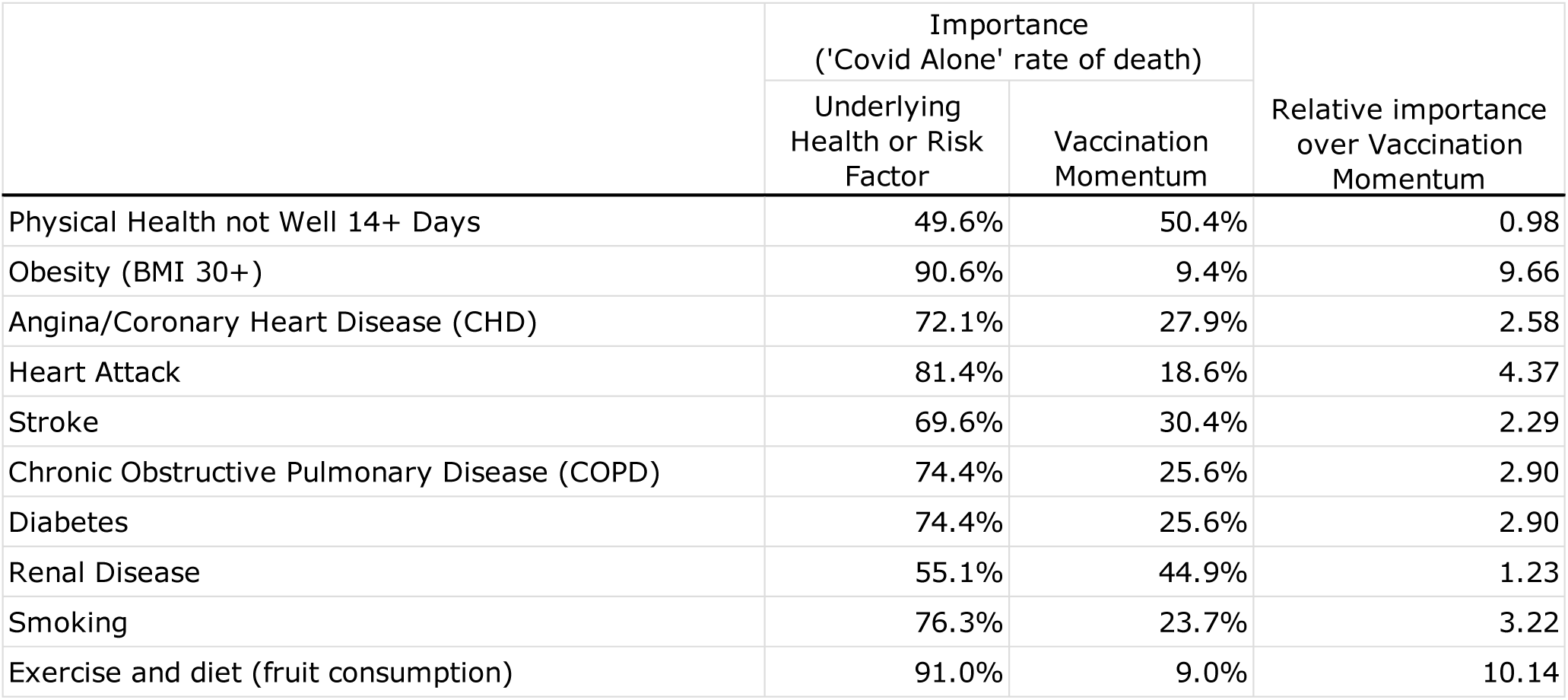
Relative importance of selected underlying health and risk factors over vaccination momentum on ‘Covid Alone’ rates of death

On 1 December 2021, the “World Health Assembly agreed to kickstart a global process to draft and negotiate a convention, agreement or other international instrument under the Constitution of the World Health Organization to strengthen pandemic prevention, preparedness and response.” [15]

According to the European Union one of the key purposes of this international agreement on pandemics is to ensure “A globally coordinated approach to discovering, developing and delivering effective and safe medical solutions, such as vaccines, medicines, diagnostics and protective equipment”. [16]

Based on the analysis presented in this paper, a singular focus on “vaccines, medicines, diagnostics and protective equipment”, *after* the next pandemic has erupted, is entirely reactive, neglects the major contributors to mitigating the effects of respiratory and other pandemics (i.e. the underlying health and wellness of national populations), and will direct already constrained funding and resources to the levers with the smaller population-level impact.

If medical and health regulators are to deliver a quantum decrease in COVID19 deaths they should be urging governments and private organisations mandate greater exercise, weight loss, less junk food, and better nutrition. And a concerted effort at reducing chronic adverse health conditions.

## Data Availability

All data produced in the present study are available upon reasonable request to the authors

